# Sleep in disorders of consciousness: Behavioral and polysomnographic recording

**DOI:** 10.1101/2020.05.21.20106807

**Authors:** Isabella Mertel, Yuri G. Pavlov, Christine Barner, Friedemann Müller, Susanne Diekelmann, Boris Kotchoubey

**Author notes:** The authors contributed equally (Isabella Mertel, Yuri G. Pavlov). Corresponding author: Yuri G. Pavlov, Address: Institute of Medical Psychology and Behavioral Neurobiology, University of Tübingen, 72076, Germany.

## Abstract

**Background:** Sleep-wakefulness cycles are an essential diagnostic criterion for Disorders of Consciousness (DOC), differentiating prolonged DOC from coma. Specific sleep features, like the presence of sleep spindles, are an important marker for the prognosis of recovery from DOC. Based on increasing evidence for a link between sleep and neuronal plasticity, understanding sleep in DOC might facilitate the development of novel methods for rehabilitation. Yet, well-controlled studies of sleep in DOC are lacking. Here, we aimed to quantify, on a reliable evaluation basis, the distribution of behavioral and neurophysiological sleep patterns in DOC over a 24h period while controlling for environmental factors (by recruiting a group of conscious tetraplegic patients who resided in the same hospital).

**Methods:** We evaluated the distribution of sleep and wakefulness by means of polysomnography (EEG, EOG, EMG) and video recordings in 32 DOC patients (16 Unresponsive Wakefulness Syndrome [UWS], 16 Minimally Conscious State [MCS]) and 10 clinical control patients with severe tetraplegia. Three independent raters scored the patients’ polysomnographic recordings.

**Results:** All but one patient (UWS) showed behavioral and electrophysiological signs of sleep. Control and MCS patients spent significantly more time in sleep during the night than during daytime, a pattern that was not evident in UWS. DOC patients (particularly UWS) exhibited less REM sleep than control patients. 44% of UWS patients and 12% of MCS patients did not have any REM sleep, while all control patients (100%) showed signs of all sleep stages and sleep spindles. Furthermore, no sleep spindles were found in 62% of UWS patients and 21% of MCS patients. In the remaining DOC patients who had spindles, their number and amplitude were significantly lower than in controls.

**Conclusions:** The distribution of sleep signs in DOC over 24 hours differs significantly from the normal sleep-wakefulness pattern. These abnormalities of sleep in DOC are independent of external factors such as severe immobility and hospital environment.

## Background

Acquired brain injury can result in a prolonged state of severe disturbance or even the lack of awareness, referred to as severe disorders of consciousness (DOC). According to the widely accepted definition [1], the presence of sleep-wakefulness cycle serves as an important symptom distinguishing DOC from acute coma. This alternation of sleep and wakefulness, however, is evaluated only at a behavioral level, that is, as the presence of episodes with open and closed eyes. More recent data indicate that, on the one hand, in some DOC patients the behavioral sleep-wakefulness cycles do not correspond to any neurophysiological signs of sleep and wakefulness [2]. On the other hand, some acute coma patients can exhibit elements of sleep such as REM-sleep, sleep spindles [3], and K-complexes [4].

Two subgroups of DOC are Unresponsive Wakefulness Syndrome (UWS), where patients have reflexive responses but no sign of awareness, and Minimally Conscious State (MCS), in which patients show unstable conscious behavior but cannot communicate or intentionally use objects. The differential diagnosis between the two is extremely challenging and error prone [5], which is additionally complicated by strong fluctuations of the arousal level and the associated level of consciousness, particularly in MCS [6], but also in UWS [7], generating variability in the results of repeated behavioral evaluations [8].

Polysomnographic recordings could contribute to the diagnostics of DOC by improving the coordination of task-based diagnostic measurements [9]. Additionally, sleep parameters such as the presence of slow wave sleep (SWS), rapid eye movements sleep (REM) and sleep spindles may serve as independent markers of the severity of consciousness impairment [10–12]. Some studies even suggest that these sleep parameters may predict the clinical outcome of DOC, i.e. whether or not patients will regain consciousness [13–16].

Although the body of literature about sleep in DOC has substantially increased over the last years, a number of serious issues remain [17]. First, there is no consensus on systematic sleep stage classification in DOC patients [12, 18–21]. The standard sleep criteria used in healthy individuals [22, 23] cannot be applied directly, but have to be adjusted to DOC sleep patterns [20]. To our best knowledge, DOC sleep data are either scored by a single rater, whose blinding is rarely warranted, or a pure automatic analysis is performed. The former substantially decreases the reliability of sleep evaluation, and the latter reduces its validity because the results of the automatic EEG assessment cannot be directly projected onto sleep stages.

Second, many polysomnographic recordings in DOC have been performed during the night (8-10 hours) or an “extended night” interval (16 hours) [2, 12, 24]. Thus, sleep of patients who slept during the day might have been incorrectly assessed.

Third, as noted above, it remains unclear whether the ‘behavioral wakefulness and sleep’, used as an important differential diagnostic sign of DOC and measured by the presence of opened/closed eyes episodes, really corresponds to ‘electrophysiological sleep’ (derived from polysomnography).

Fourth, none of the studies so far included clinical control groups but only healthy controls. Yet, the living conditions of severely disabled DOC patients radically differ from that of healthy individuals, regarding many parameters that can affect sleep quality, such as the ward room, immobility, the lack of social pressure and unsolicited disturbance by the hospital personnel.

To solve these issues, in this study we carried out reliable expert evaluation of behavioral and electrophysiological sleep over a 24h period in two groups of DOC patients (UWS and MSC) and a group of tetraplegic control patients.

## Methods

### Patients

The study was conducted in accordance with the ethical standards of the Declaration of Helsinki and was approved by the Ethics Committee of the Faculty of Medicine, University of Tübingen. The patients and their legal representatives were informed about the study content before study enrollment and gave their written consent. They were informed that participation in the study had no effect on medical treatment and that they can terminate their participation at any time without negative consequences. The study was registered in the German Clinical Study Register (DRKS00009326).

Patients were included according to the following criteria: age between 18-69 years (to minimize age-dependent effects on sleep [25]); time post ictum from 1-24 months; stable circulation and respiration. Exclusion criteria were: basic EEG activity below 3 Hz; a history of depression, schizophrenia, drug abuse, sleep disorders, or any neurological disorders; epileptic activity in premorbid EEG recordings. We did not explicitly include extremely severe background EEG patterns (such as isoelectric EEG or burst-suppression pattern) in the list of exclusion criteria because we had not observed such patterns among hospitalized DOC patients in our earlier samples [e.g., 26, 27]. These patterns were not observed in the present sample either. It should be noted that DOC patients in rehabilitation hospitals are not necessarily representative for the entire DOC population. Nevertheless, if patients with these patterns were found in the current study, they would have been excluded. The diagnoses of DOC patients were either UWS or MCS. The diagnoses of Clinical Control group patients (CC) were Guillain-Barré syndrome or severe high-level spinal cord injury with tetraplegia. Patients were included in the CC group only if their time out of bed did not exceed 4 h/day.

All patients admitted to the Schön Clinics Bad Aibling (Bavaria, Germany) in the period from February 2016 to February 2018 who fulfilled the above criteria were initially enrolled in the study, resulting in a sample of nineteen UWS, 19 MCS, and 12 CC patients. However, two control patients withdrew their consent. Further six DOC patients were excluded, because their condition worsened (n = 3) or improved (n = 3) and they did not meet the inclusion criteria anymore. Therefore, data from sixteen UWS, 16 MCS, and 10 CC patients were recorded and analyzed (see Table 1).

**Table 1.** Clinical details of patients

| *id* | *Age range*<br>*(y)* | *Sex* | *Time (since*<br>*injury(d))* | *Medication*<br>*(Sed, Stim,*<br>*Anticon)* | *Etiology* | *EEG*<br>*grade* | *CRS-r*<br>*(A,V,M, OV,C,Ar)* |
| --- | --- | --- | --- | --- | --- | --- | --- |
| *UWS1* | 51-55 | m | 62 | 0, 0, 1 | hypox | 1 | 5 (1,0,2,1,0,1) |
| *UWS2* | 46-50 | m | 151 | 1, 0, 0 | TBI | 2 | 5 (1,0,1,1,0,2) |
| *UWS3* | 61-65 | f | 273 | 0, 0, 0 | stroke | 5 | 7 (2,1,2,1,0,1) |
| *UWS4* | 26-30 | f | 68 | 0, 0, 1 | TBI | 3 | 7 (2,1,1,1,0,2) |
| *UWS5* | 51-55 | f | 134 | 1, 0, 1 | hypox | 1 | 5 (1,0,1,1,0,2) |
| *UWS6* | 36-40 | f | 69 | 0, 1, 1 | stroke | 1 | 4 (1,0,0,1,0,2) |
| *UWS7* | 46-50 | m | 174 | 2, 0, 1 | TBI | 2 | 8 (2,1,2,1,0,2) |
| *UWS8* | 26-30 | f | 233 | 1, 0, 1 | TBI | 2 | 4 (1,0,1,1,0,1) |
| *UWS9* | 56-60 | m | 46 | 0, 0, 0 | hypox | 3 | 7 (2,1,2,1,0,1) |
| *UWS10* | 61-65 | f | 197 | 1, 1, 1 | SAH | 4 | 6 (1,0,2,1,0,2) |
| *UWS11* | 18-25 | f | 134 | 0, 0, 1 | TBI | 3 | 2 (0,0,0,1,0,1) |
| *UWS12* | 41-45 | f | 52 | 0, 0, 2 | hypox | 3 | 5 (1,0,1,2,0,1) |
| *UWS13* | 56-60 | m | 98 | 3, 0, 1 | hypox | 1 | 6 (2,0,2,1,0,1) |
| *UWS14* | 56-60 | m | 116 | 1, 1, 0 | stroke | 3 | 3 (0,0,2,1,0,0) |
| *UWS15* | 18-25 | m | 169 | 0, 1, 0 | TBI | 3 | 6 (2,0,1,1,0,2) |
| *UWS16* | 56-60 | f | 61 | 0, 0, 1 | stroke | 3 | 7 (2,0,2,1,0,2) |
| *MCS1* | 66-70 | f | 69 | 1, 0, 1 | stroke | 4 | 10 (2,2,2,1,1,2) |
| *MCS2* | 56-60 | m | 155 | 0, 0, 1 | hypox | 2 | 12 (2,1,5,2,1,1) |
| *MCS3* | 51-55 | m | 84 | 1, 1, 1 | hypox | 1 | 9 (2,0,2,2,1,2) |
| *MCS4* | 46-50 | m | 109 | 0, 2, 1 | TBI | 4 | 10 (2,3,2,1,0,2) |
| *MCS5* | 51-55 | m | 60 | 2, 0, 0 | TBI | 5 | 14 (3,3,5,1,1,1) |
| *MCS6* | 51-55 | f | 158 | 1, 1, 1 | SAH | 4 | 12 (2,3,2,2,1,2) |
| *MCS7* | 61-65 | m | 66 | 0, 1, 1 | hypox | 1 | 6 (2,2,0,1,0,1) |
| *MCS8* | 61-65 | m | 133 | 0, 1, 0 | SAH | 3 | 9 (2,3,2,1,0,1) |
| *MCS9* | 21-25 | m | 150 | 0, 1, 0 | hypox | 4 | 9 (1,0,5,1,0,2) |
| *MCS10* | 66-70 | m | 140 | 0, 1, 0 | TBI | 4 | 16 (2,3,5,3,1,2) |
| *MCS11* | 21-25 | f | 234 | 3, 0, 0 | enceph | 2 | 8 (0,1,4,1,0,2) |
| *MCS12* | 51-55 | m | 92 | 0, 0, 0 | SAH | 5 | 11 (2,3,2,2,0,2) |
| *MCS13* | 18-25 | f | 131 | 1, 0, 0 | stroke | 3 | 8 (1,0,5,1,0,1) |
| *MCS14* | 41-45 | m | 78 | 0, 1, 1 | SAH | 4 | 8 (1,2,2,1,0,2) |
| *MCS15* | 41-45 | m | 96 | 1, 1, 1 | hypox | 1 | 9 (2,2,2,1,0,2) |
| *MCS16* | 56-60 | f | 74 | 1, 0, 1 | stroke | 5 | 14 (3,1,5,2,1,2) |
| *CC1* | 51-55 | m | 155 | 1, 0, 0 | - | - | - |
| *CC2* | 26-30 | f | 1176 | 1, 0, 2 | - | - | - |
| *CC3* | 36-40 | m | 3876 | 1, 0, 1 | - | - | - |
| *CC4* | 18-25 | m | 379 | 1, 0, 1 | - | - | - |
| *CC5* | 65-70 | f | 33 | 0, 0, 0 | - | - | - |
| *CC6* | 61-65 | m | 267 | 1, 0, 1 | - | - | - |
| *CC7* | 51-55 | f | 131 | 1, 0, 0 | - | - | - |
| *CC8* | 66-70 | m | 96 | 1, 0, 1 | - | - | - |
| *CC9* | 26-30 | m | 682 | 0, 0, 0 | - | - | - |
| *CC10* | 18-25 | m | 163 | 1, 0, 0 | - | - | - |

Notes: CRS-r: total score of Coma Recovery Scale – revised, A auditory, V visual, M motor, OV oromotor/verbal, C communication, Ar arousal subscores; Etiology: TBI – traumatic brain injury, hypox – hypoxic brain injury, enceph – encephalitis, SAH – subarachnoid hemorrhage.

A trained and experienced neurologist repeatedly performed clinical assessment of DOC patients using the Coma Recovery Scale – revised (CRS-r [28]). The last CRS-r evaluation was done on the day before the polysomnographic recording (see Table 1). UWS and MCS groups did not differ in terms of age (UWS: 46.8 ± 14.6 years; MCS: 48.8 ± 14.8 years), gender (UWS: m/f = 7/9; MCS: m/f = 11/5), time since injury (127 ± 68.6 days and 114 ± 46.6 days, for UWS and MCS, respectively), as well as the type of injury (traumatic/non-traumatic ratio 6/10 and 3/13, for UWS and MCS, respectively). In respect of age and gender, DOC patients also did not significantly differ from clinical control group patients (m/f = 7/3, age 43.7 ± 18.5 years).

### Recording

A continuous 24h polysomnographic recording included EEG, two channels of electrooculography (EOG), with the electrodes positioned 1 cm lateral and below and above to the outer canthi of both eyes, three channels of electromyography (EMG), with the electrodes placed on the chin, and video recording. Single cups EEG electrodes were attached at F3, F4, Fz, C3, C4, Cz, P3, P4, Pz and both mastoids, referenced to Cz with the ground electrode at Fpz.

The data were recorded using a mobile polysomnography device (SomnoScreenplus, Somnomedics, Germany) with 256 Hz sampling rate with 0.2 Hz high-pass and 35 Hz low-pass filters.

All recordings were performed in the patients’ wards on weekends when no rehabilitative treatments took place. During the recording, all patients (including control patients) had to be moved by nurse personnel to prevent decubitus. After each such intervention, the experimenter checked electrode impedance and renewed electrodes whenever the impedance exceeded 5 kΩ.

The individual medication intake had remained stable during at least one week before recording. Recordings started between around 10 am and 12 noon and ended at the same time on the next day.

### Data analysis

Behavioral sleep-wakefulness state of each patient was defined in 30-second intervals based on video recordings. An interval was classified as sleep if the eyes were closed at the onset of the interval, and as wakefulness, if they were open.

Before the evaluation of electrophysiological sleep-wakefulness state, EEG data were re-referenced to averaged mastoids and pre-processed including a notch filter (50 Hz). The program SchlafAus (developed by Steffen Gais, unpublished) was used for sleep scoring in 30-second intervals. Overall, there are 2880 30-s intervals during 24 hours, but the number of assessments per recording varied because sometimes more than 24 hours were recorded, and some epochs were lost due to the amplifier’s battery replacement (mean number of epochs = 2868, SD = 30.5, range 2740-2946).

Three independent raters (IM, YGP, CB) scored the data under the supervision of the most experienced fourth person (SD). All raters had previous experience in the classification of sleep stages and were familiar with the essential characteristics of patient groups. Owing to the randomization of patient numbers, all raters were blind to clinical and demographic patient data.

The first ten recordings were scored by all three raters independently of each other. After having rated two or three patients, they met and discussed their agreements and disagreements. The supervisor defined the main points of discordance to be discussed. At the end of each meeting scoring criteria, originally based on those of Rechtschaffen and Kales [22], were adapted to maximize a pairwise concordance in each of the three pairs of raters.

After the first ten patients, no further discussion was allowed anymore. The following 32 recordings were randomly distributed among the three pairs of raters. Within each pair, each rater worked independently of the other one. The average percentage agreement (i.e., the number of 30-second epochs on which two raters completely agreed, divided by the total number of epochs evaluated by these two raters and multiplied by 100) for the first ten cases was 83.53 ± 3.84%, and for subsequent cases 82.23 ± 8.06%. In case of conflicting scorings, the raters agreed on the final version where all scoring conflicts were resolved by mutual consensus on an epoch-by-epoch basis.

The modifications of scoring criteria, as compared with Rechtschaffen and Kales [22], were as follows. Firstly, DOC patients may show local intermittent rhythmic delta activity that might be confused with sleep-related delta waves. For this reason, nine rather than two EEG electrodes were used for sleep scoring. This permitted us to detect deviating channels and exclude them from classification.

Secondly, since the EEG amplitude in patients can be influenced by many extracerebral factors, the amplitude criterion for slow waves (>75 µV) was not followed. Thirdly, some patients did not show typical markers for sleep stage S2 (sleep spindles, K complexes). In such cases S2 was assessed on the basis of EEG slowing, the appearance of slow wave activity), and a further reduction of muscle tone. Slow waves, however, should not cover more than 20% of the epoch, otherwise S3 or S4 was scored.

Sleep spindle activity can be visually evaluated, automatically detected, or deducted from the EEG spectrum [29, 30]. In the literature there is no agreement on which approach is to be preferred [31–33]. In the present study we applied both methods: a visual identification of spindles by two independent scorers and, then, an automatic algorithm (with default settings) for sleep spindle quantification [34] to artifact-free stage S2 epochs at Cz channel. The following statistical analyses involved only patients with at least one clearly present visually detected sleep spindle in the recording. However, Table 2 shows spindle data (frequency, density, and amplitude) according to the automatic spindle detection in all patients.

**Table 2.** Sleep characteristics

| *id* | *S1, min* | *S2, min* | *SWS, min* | *REM, min* | *Spindles present* | *N spindles* | *Density, sp/min* | *Amplitude, $\mu V$* |
| --- | --- | --- | --- | --- | --- | --- | --- | --- |
| *UWS1* | 88 | 389.5 | 22 | 4.5 | 0 | 19 | 0.06 | 19.57 |
| *UWS2* | 33.5 | 434.5 | 31 | 5.5 | 0 | 31 | 0.08 | 14.28 |
| *UWS3* | 68.5 | 194 | 86 | 0 | 0 | 0 | 0 | 0 |
| *UWS4* | 15.5 | 77.5 | 72.5 | 7.5 | 0 | 0 | 0 | 0 |
| *UWS5* | 171 | 189 | 27 | 28.5 | 0 | 10 | 0.06 | 6.86 |
| *UWS6* | 0 | 0 | 0 | 0 | 0 | 0 | 0 | 0 |
| *UWS7* | 72.5 | 77.5 | 2 | 0 | 0 | 20 | 0.26 | 11.16 |
| *UWS8* | 31 | 154 | 242 | 30.5 | 1 | 14 | 0.10 | 9.22 |
| *UWS9* | 64 | 258 | 102.5 | 30.5 | 1 | 2 | 0.01 | 7.79 |
| *UWS10* | 71 | 3 | 0 | 0 | 0 | 0 | 0 | 0 |
| *UWS11* | 2 | 479.5 | 200.5 | 0 | 1 | 89 | 0.21 | 38.29 |
| *UWS12* | 46.5 | 190.5 | 52 | 0 | 1 | 125 | 0.68 | 7.13 |
| *UWS13* | 49 | 69.5 | 2 | 1.5 | 1 | 7 | 0.15 | 6.99 |
| *UWS14* | 81 | 154 | 24.5 | 14 | 0 | 3 | 0.02 | 6.99 |
| *UWS15* | 24.5 | 64.5 | 68 | 67.5 | 1 | 65 | 1.13 | 13.62 |
| *UWS16* | 13.5 | 78.5 | 204.5 | 0 | 0 | 31 | 0.43 | 15.25 |
| UWS (M±SD) | 52±42.5 | 175.8±147.5 | 71±78.8 | 11.9±18.8 | | | | |
| *MCS1* | 83.5 | 163.5 | 33 | 0 | 0 | 1 | 0.01 | 7.80 |
| *MCS2* | 159.5 | 63 | 0 | 86.5 | 0 | 15 | 0.39 | 29.10 |
| *MCS3* | 162 | 119.5 | 47 | 7.5 | 0 | 2 | 0.02 | 7.80 |
| *MCS4* | 46.5 | 187 | 57 | 19.5 | 0 | 6 | 0.04 | 10.20 |
| *MCS5* | 119 | 88.5 | 4 | 24.5 | 1 | 143 | 1.71 | 14.01 |
| *MCS6* | 147.5 | 504.5 | 173 | 126 | 1 | 3 | 0.01 | 10.45 |
| *MCS7* | 216.5 | 110.5 | 11.5 | 28 | 1 | 8 | 0.08 | 11.71 |
| *MCS8* | 67.5 | 373.5 | 63.5 | 10.5 | 1 | 4 | 0.01 | 10.71 |
| *MCS9* | 93 | 262 | 75 | 38 | 1 | 120 | 0.48 | 18.64 |
| *MCS10* | 75 | 323.5 | 47.5 | 24 | 0 | 5 | 0.03 | 8.32 |
| *MCS11* | 4.5 | 295.5 | 253.5 | 90 | 1 | 121 | 0.61 | 8.48 |
| *MCS12* | 169 | 104 | 15 | 22 | 1 | 2 | 0.03 | 15.71 |
| *MCS13* | 25.5 | 206 | 119.5 | 0 | 1 | 19 | 0.10 | 12.21 |
| *MCS14* | 21.5 | 177 | 19.5 | 6 | 1 | 1 | 0.01 | 4.80 |
| *MCS15* | 197.5 | 225 | 6.5 | 31.5 | 1 | 323 | 1.48 | 16.48 |
| *MCS16* | 34.5 | 194.5 | 0 | 25 | 1 | 11 | 0.06 | 6.60 |
| MCS (M±SD) | 101.4±67.2 | 212.3±117.3 | 57.8±70.3 | 33.7±36 | | | | |
| *CC1* | 60 | 231 | 162.5 | 21 | 1 | 209 | 0.99 | 25.37 |
| *CC2* | 40 | 191.5 | 144 | 91.5 | 1 | 50 | 0.27 | 25.48 |
| *CC3* | 64.5 | 244.5 | 102 | 83 | 1 | 146 | 0.70 | 20.47 |
| *CC4* | 152 | 446.5 | 41 | 49.5 | 1 | 264 | 0.65 | 16.86 |
| *CC5* | 111 | 231 | 8.5 | 41 | 1 | 89 | 0.47 | 15.67 |
| *CC6* | 56.5 | 209 | 30.5 | 33 | 1 | 20 | 0.11 | 11.42 |
| *CC7* | 68.5 | 282.5 | 56 | 17 | 1 | 276 | 1.08 | 15.48 |
| *CC8* | 56 | 286 | 24.5 | 84 | 1 | 130 | 0.52 | 15.77 |
| *CC9* | 170.5 | 170.5 | 101 | 88.5 | 1 | 82 | 0.59 | 20.17 |
| *CC10* | 36.5 | 193.5 | 110.5 | 51 | 1 | 281 | 1.53 | 20.50 |
| CC (M±SD) | 81.5±46.8 | 248.6±79.1 | 78±53.2 | 56±28.7 |  |  |  |  |

Notes: UWS, unresponsive Wakefulness Syndrome; MCS, Minimally Conscious State; CC, Clinical Control; S1, S2, sleep stages 1 and 2, resp; SWS, Slow-Wave Sleep; REM, Rapid Eye Movement Sleep; Spindles present (defined by visual screening): 0 – no, 1 – yes; N Spindles, overall number of spindles; Density, sleep spindle density defined as overall number of spindles divided by the duration of artifact free S2 in minutes; Amplitude, the average amplitude of sleep spindles in $\mu V$ .

Patterns of routine wakefulness EEG were first subdivided into five classes, described in Table 1. Because the corresponding patient subgroups were too small, for statistical analyses classes 1 and 2 were combined into a category of EEG without a clear background rhythmic activity (“bad EEG”), and classes 3, 4, and 5 built together a category of EEG with well pronounced theta (or low alpha) oscillations (“good EEG”). Similarly, one can see in Table 1 that some etiological groups were small (e.g., only one patient with inflammatory etiology). Therefore, we grouped all etiologies into three categories: traumatic, hypoxic, and other.

Statistical analyses were performed using R v.3.6.0. The 24h recording was subdivided into day (08.00 - 20.00) and night (20.00 - 08.00), according to the time of intensive therapy and lights on versus the time of rest and lights off. A mixed ANOVA included a repeated measures factor DayTime (i.e., day/night) and a between-subject factor Group (UWS, MCS, CC). A similar ANOVA was also performed with the factor Etiology (3 levels: traumatic, hypoxic, other; CC patients excluded). Kruskal-Wallis and Wilcoxon Rank Sum tests were used for comparing proportions between the groups. When appropriate, we tested the lack of difference between the groups by calculating Bayes Factors (BF) using a Bayesian ANOVA or *t*-test. For this analysis, we used BayesFactor package for R with default priors.

## Results

### Behavioral and electrophysiological sleep

The overall proportion of eyes-closed and eyes-opened epochs in the 24h period was close to 50/50% (UWS: 49%, MSC: 51%, CC: 47% eyes-closed epochs) and did not differ between all three groups (*p* = .89, BF_01_ = 200). Interestingly, the groups differed in the proportion of open and closed eyes epochs during night and daytime (DayTime x Group: *F*(2, 39) = 8.11, *p* = .001, η^2^= .29). While CC (*t*(9) = 4.21, *p* = .0023, *d* = 1.40) and MCS patients (*t*(15) = 2.41, *p* = .03, *d* = 0.62) spent a greater amount of time with closed eyes during the night than daytime, this pattern was not evident in UWS patients (*t*(15) = 0.35, *p* = .73, *d* = 0.09, BF_01_ = 3.7; see Figure 1A).

**Figure 1.**
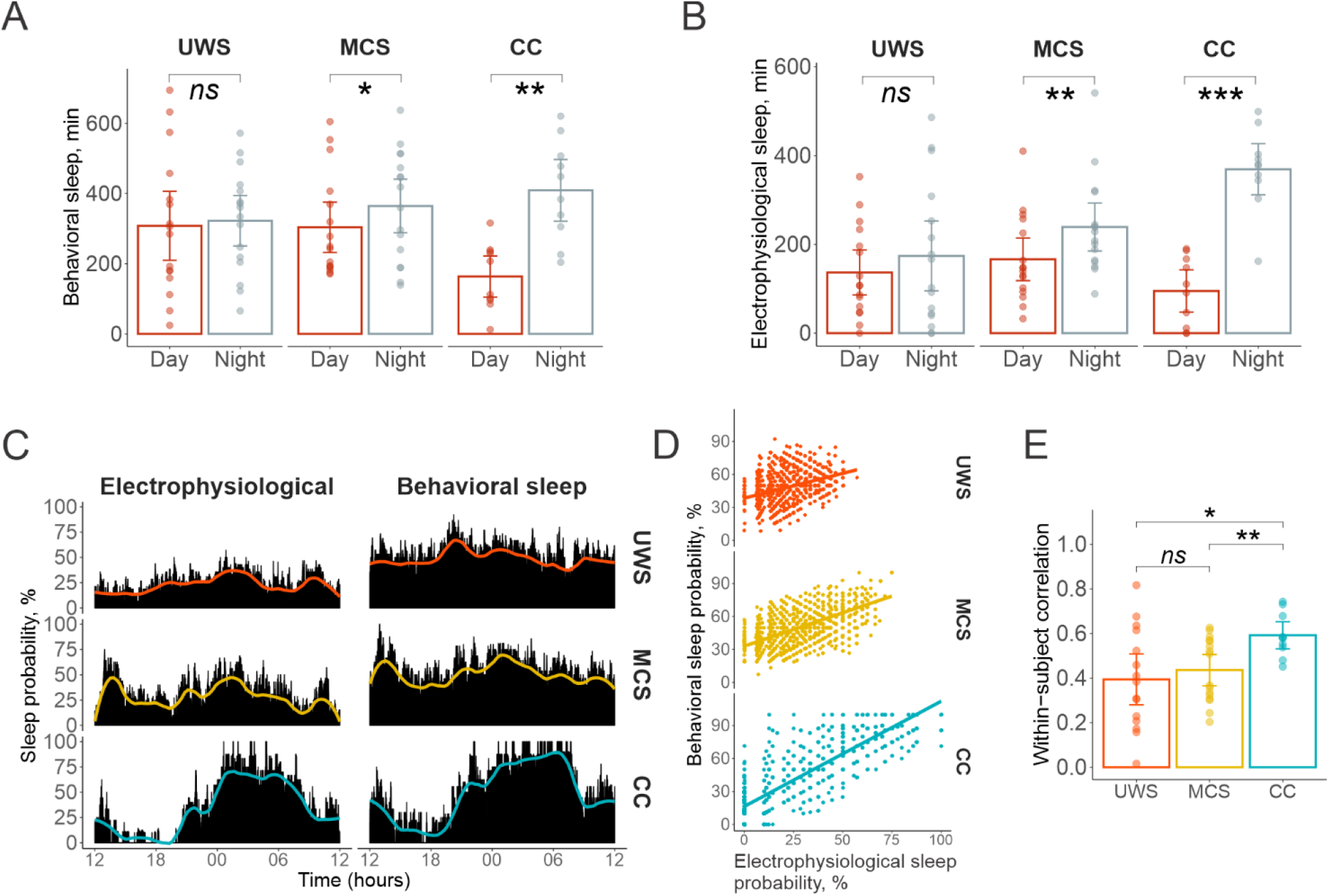
Behavioral and electrophysiological sleep. UWS, Unresponsive Wakefulness Syndrome; MCS, Minimally Conscious State; CC, Clinical Control. (A) The amount of behavioral sleep (number of epochs with eyes closed) during the night and at daytime, for each group. (B) The amount of electrophysiological sleep evaluated by polysomnographic recording during the night and at daytime, for each group. (C) The distribution of sleep probability (percentage of patients who slept during the respective epoch) across all 2880 epochs. Solid lines show the results of smoothing according to the LOESS algorithm with the smoothing span of 0.2 by means of ggplot2 R package. (D) Scatterplots relating the probabilities (in %) that a particular epoch was a sleep epoch as scored with electrophysiological and behavioral measures (each dot represents one epoch). Note that the whole graph is “larger” for CC than for UWS and MCS, indicating a higher behavioral-electrophysiological correspondence among CC patients than DOC patients. There were epochs when all CC patients slept, and epochs when all of them where awake, but there were no such epochs in the two DOC groups. (E) Behavioral-electrophysiological sleep Kendall correlations calculated within each subject (dots) across epochs and averaged for each group (columns). * p < .05, ** p < .01, *** p < .001, *^ns^* not significant. Error bars are 95% confidence intervals.

When electrophysiological sleep data were dichotomized (i.e., 1 – any stage of sleep, 0 – wakefulness), the same pattern was observed. CC (*t*(9) = 6.21, *p* = .0002, *d* = 2.07) and MCS (*t*(15) = 3.04, *p* = .008, d = 0.79) but not UWS (*t*(15) = 0.76, *p* = .46, *d* = 0.20, BF_01_ = 3) patients slept longer during the night than during the day (DayTime x Group: *F*(2, 39) = 8.19, *p* = .001, η^2^ = .30, see Figure 1B). Eight of the ten CC patients took a daytime nap. One UWS patient did not show any signs of electrophysiological sleep despite episodes of closed eyes. All other DOC patients had at least a short interval of sleep during daytime. One UWS patient slept only during daytime. Figure 2 shows examples of individual patients’ hypnograms.

**Figure 2.**
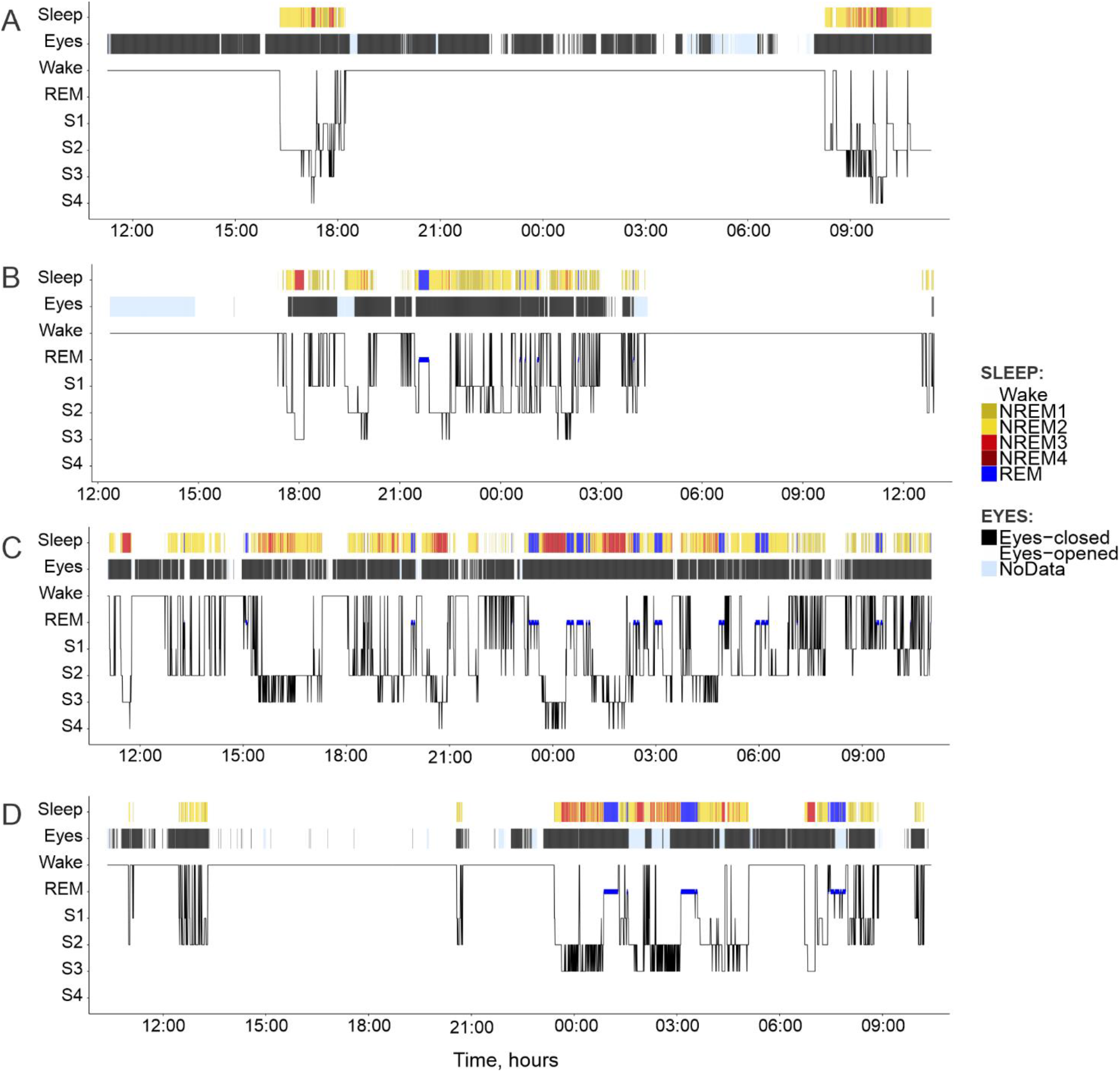
Exemplary hypnograms of four patients. (A) an UWS patient who remained awake at night but slept during the day; (B) an MCS patient with close-to-normal sleep distribution; (C) an UWS patient with uniformly distributed sleep over the 24h period; (D) a CC patient with a pattern of well-structured sleep during the night and an afternoon nap. Notes: Sleep – electrophysiological sleep, Eyes – behavioral sleep.

Sleep of DOC patients appeared to be characterized by frequent changes between sleep and wakefulness. To check this impression, we calculated the number of transitions between sleep and wakefulness (i.e., the intervals that were classified as different from the immediately preceding intervals). The number of transitions did not significantly differ among groups, neither on the basis of behavioral sleep data (*F*(2, 39) = 1.09, *p* = .35, η^2^ =.05) nor when the electrophysiological data were used (F(2, 38) = 1.46, *p* = .24, η^2^ =.24). The number of transitions in the electrophysiological sleep data analysis was corrected for the number of sleep epochs because of unequal sleep duration between the patients. One UWS patient who did not sleep at all was excluded from this analysis.

To examine the correspondence between behavioral and electrophysiological data, we calculated the percentage of patients who slept in each of the 2880 epochs (sleep probability). Rank-order correlations between sleep probabilities based on behavioral and electrophysiological data were highly significant in all three groups (*rho* = 0.87, 0.58, and 0.37 for CC, MCS, and UWS, respectively; all *ps* < 0.001). All three correlations were pairwise significantly different from each other, with all *ps* < 0.001. Likewise, the correspondence between behavioral and electrophysiological sleep differed between groups when it was analyzed on the basis of single epoch data within each patient (*F*(2, 38) = 4.27, *p* = .021, η^2^ = .18).

### Sleep stage distribution

As shown in Figure 3, the total amount of sleep as measured with polysomnographic recordings did not significantly differ between the groups (*F*(2, 39) = 2.76, *p* = .08, η^2^ = .12), although post-hoc tests revealed shorter overall sleep duration in UWS patients compared to the CC group. With regard to single sleep stages, there were significant group differences concerning the time spent in sleep stage S1 (*F*(2, 39) = 3.36, *p* = .045, η^2^ = .15, BF_10_ = 1.8) and REM sleep (*F*(2, 39) = 7.41, *p* = .002, η^2^ = .28, BF_10_ = 19.4), but not in S2 (*F*(2, 39) = 1.1, *p* = .34, η^2^ = .05, BF_01_ = 2.73) and SWS (*F*(2, 39) = 0.28, *p* = .75, η^2^ = .01, BF_01_ = 4.71). MCS patients spent more time in S1 than UWS patients (t(30) = 2.49, p = .019, d = 0.88). Further, UWS patients spent less time in REM sleep than CC patients (t(24) = 4.76, *p* < .001, *d* = 1.92) and MCS (t(30) = 2.15, p = .040, d = 0.76).

**Figure 3.**
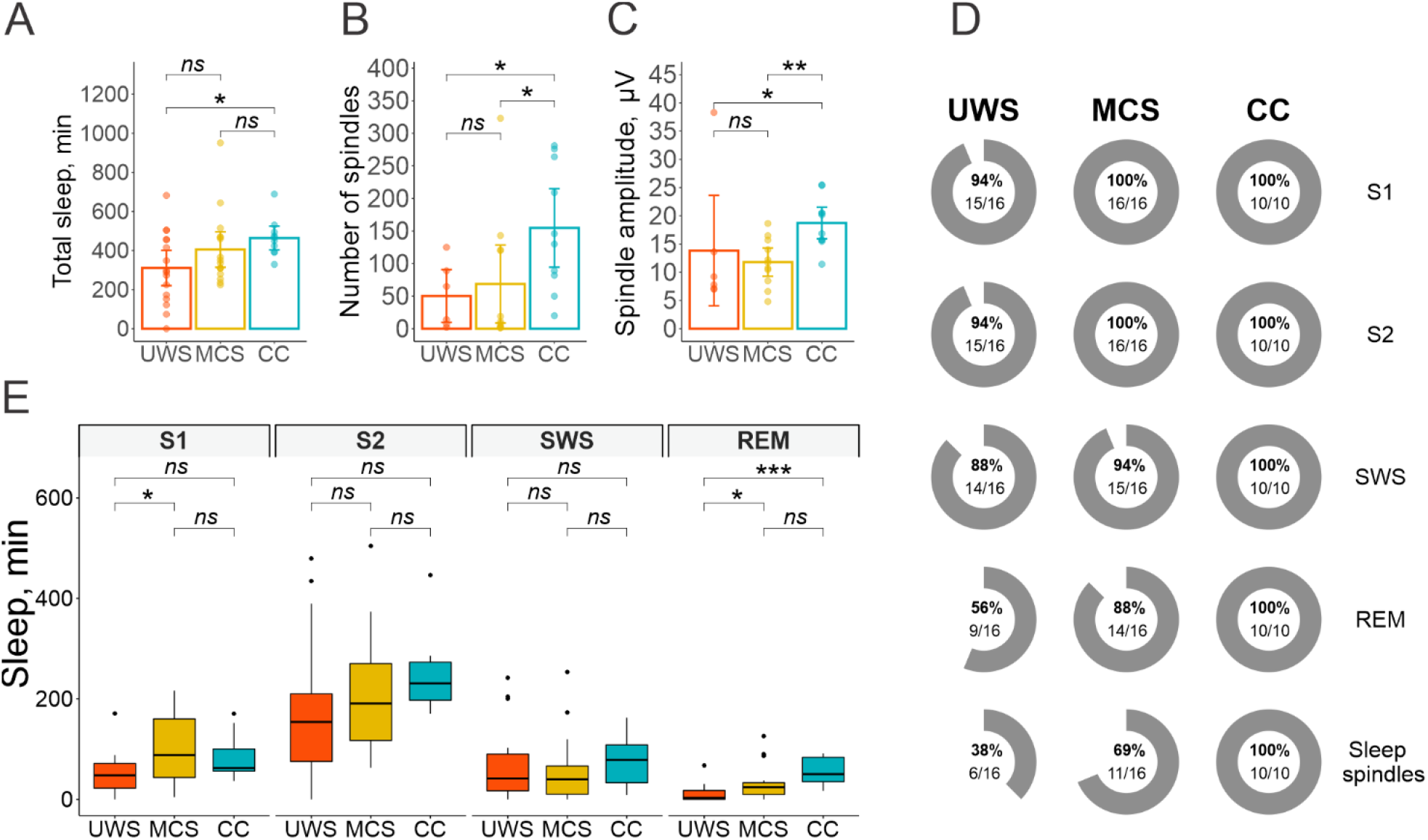
Sleep stage distribution. (A) Total amount of sleep by group. Mean ± SD: 311 ± 184, 405 ± 185, 464 ± 98 min in UWS, MCS and CC groups respectively. (B) Number of sleep spindles by group. Mean ± SD: 50.3 ± 50.6, 68.6 ± 101, 155 ± 97.2 spindles in S2 in UWS, MCS and CC groups respectively. Only patients with present spindles (defined by visual screening) included (see Table 2). (C) Amplitude of sleep spindles by group. Mean ± SD: 13.8 ± 12.2, 11.8 ± 4.23, 18.7 ± 4.52 µV in UWS, MCS and CC groups respectively. Only patients with present spindles (defined by visual screening) included (see Table 2). (D) The percentage of patients in each group, showing signs of the respective sleep stage during the 24h recording period. (E) Time spent in single sleep stages by group. UWS, unresponsive Wakefulness Syndrome; MCS, Minimally Conscious State; CC, Clinical Control; SWS, Slow-Wave Sleep; REM, Rapid Eye Movement Sleep; S1, S2, sleep stages 1 and 2, resp.; * *p* < .05, ** *p* < .01, *** *p* < .001, *ns* not significant. Error bars are 95% confidence intervals.

As depicted in Figure 3D, sleep stages S1, S2, SWS, and REM sleep were present in all CC patients, but not in all UWS and MCS patients. Statistically, the REM stage was less frequently found in UWS than in MCS (*χ*^2^(1) = 3.74, *p* = .053) and in CC (*χ*^2^(1) = 5.76, *p* = .016) but MCS did not differ from CC (*χ*^2^(1) = 1.3, *p* = .25), resulting in a generally significant Group effect (*χ*^2^(2) = 8.02, *p* = .018). All other sleep stages did not differentiate the groups significantly.

When diagnostic groups (UWS, MCS) were replaced with etiological groups (traumatic, hypoxic, other; CC patients excluded), hypoxic patients were found to sleep significantly longer in stage S1 (mean = 124.7 min) than non-hypoxic patients (means = 46.6 min and 60.6 min for TBI and other etiologies, respectively; *F*(2, 29) = 6.28, *p* = .005, η^2^ = .30). Other sleep stages were not significantly related to etiology (S2: *F*(2, 29) = 0.08, *p* = .92, η^2^ < .01; SWS: *F*(2, 29) = 1.22, *p* = .31, η^2^ = .08; REM: *F*(2, 29) = 0.08, *p* = .92, η^2^ < .01). Likewise, we were unable to find any relationship of sleep data with morphological characteristics of brain lesions, possibly because of the difficulty with lesion classification and grouping (see Additional file 1: Table S1).

### Effects of age and background EEG

As it is known that the proportion of SWS and REM sleep decrease with age [25], we tested the possibility that our results were affected by age-related changes in sleep architecture. We confirmed general SWS decrement with age in our sample (Spearman correlation between the amount of SWS and age in years: *rho* = −0.44, p = .004). A correlation of smaller magnitude with age was found for S1 (*rho* = 0.34, p = .027) but not for REM sleep (*rho* = −0.18, p = .25) or S2 (*rho* = −0.1, p = .53). When introducing age as a covariate in our overall sleep stage analysis, the effect of Age was only significant for SWS (*F*(1, 38) = 10.75, *p* = .002, η^2^ = .22). Controlling for age made the effect of Group on SWS even weaker (*F*(2, 38) = 0.1, *p* = .90, η^2^ = .005, BF_01_ = 5.29), while producing negligible effects on REM sleep (*F*(2, 38) = 7.33, *p* = .002, η^2^ = .28), S1 (*F*(2, 38) = 3.34, *p* = .05, η^2^ = .15), and S2 (*F*(2, 38) = 1.02, *p* = .37, η^2^ = .05), suggesting that the observed differences between patient groups were not considerably affected by age.

We further checked whether the background EEG activity, obtained by standard clinical EEG recordings, was related to sleep. None of the sleep stages significantly differed between patients with “good” versus “bad” EEG (S1: *F*(1, 30) = 2.67, *p* = .113, η^2^ = .08; S2: *F*(1, 30) = 0.30, *p* = .587, η^2^ < .01; SWS: *F*(1, 30) = 0.40, *p* = .533, η^2^ = .01; REM: *F*(1, 30) = 0.23, *p* = .632, η^2^ < .01). On the other hand, the background EEG grade significantly depended on etiology (p = 0.021, Fisher’s exact test). “Bad” EEG characterized seven of the 10 hypoxic patients (70%), but only three of the 9 TBI patients (33%) and two of the 13 patients with vascular and other etiologies (15%).

### Sleep spindles

Table 2 and Figure 3D show that the characteristic sleep spindles in S2 were observed in all CC patients but only in a portion of DOC patients, yielding a significant Group effect (*χ*^2^(2) = 10.44, *p* = .005). Specifically, fewer UWS patients (38%) showed any sleep spindles than CC patients (100%) (*χ*^2^(1) = 9.76, *p* = .002); the proportion of MCS patients with identified spindles (69%) only marginally differed from CC patients (*χ*^2^(1) = 3.04, *p* = .08) and UWS patients (*χ*^2^(1) = 3.72, *p* = .054).

To further explore the quality of sleep spindles, only patients with sleep spindles were included, i.e. *n*(UWS) = 6, *n*(MCS) = 11, *n*(CC) = 10. Although the average number of spindles significantly differed between the groups (*χ*^2^(2) = 6.95, *p* = .03), the difference in sleep spindle density did not reach significance (CC: 0.689 ± 0.415, MCS: 0.415 ± 0.619, UWS: 0.382 ± 0.436 spindles/min; *χ*^2^(2) = 4.6, *p* = .1). CC patients showed a significantly higher number of spindles in sleep stage S2 than UWS patients (U = 87, *p* = .024) and MCS patients (U = 50.5, *p* = .029), whereas the MCS and UWS groups did not differ (U = 31.5, *p* = .919). Similarly, UWS and MCS patients had a lower average spindle amplitude than CC patients (*U* = 49, *p* = .042 and *U* = 95, *p* = .004 respectively), yielding a significant overall group effect (*χ*^2^(2) = 8.85, *p* = .012).

## Discussion

Previous studies indicated that sleep evaluations are a promising tool in the assessment of DOC patients as the presence of sleep stages seems to be related to the severity of consciousness impairment [10, 11, 13, 35]. Sleep markers can have a higher prognostic value in DOC than many other neurophysiological tests, including event related brain potentials (ERP) and functional magnetic resonance imaging (fMRI) [14, 36]. Other authors propose a combination of ERP and polysomnography as a predictive tool in DOC [21]. Compared with many other neurophysiological methods, polysomnographic recordings are cost-effective and do not require an immediate response to a stimulus and are, therefore, independent of patient comorbidities (e.g. sensory deficits) and abnormal latency of brain responses.

A number of factors decrease the reliability of previous sleep studies in DOC. Due to numerous abnormalities of EEG and sleep patterns, sleep scoring in DOC is highly challenging. Oscillations in the same frequency range (e.g., 2-3 Hz) can indicate both normal SWS and severe brain pathology of DOC patients. Therefore, the mere description of spectral properties and spectrum-based automatic analyzes provide limited insights. The problem is further worsened by the lack of concordant data from independent, blinded raters. In the present study, a group of experienced sleep raters was explicitly trained to adjust Rechtschaffen & Kales’ [22] scoring criteria to the particular features of EEG in DOC. Each dataset was independently scored by two raters, who were unaware of the patient’s clinical data. The between-rater agreement above 80% can be regarded as high even for normal sleep [37].

In agreement with previous studies [9, 24, 38, 39], severe abnormalities of circadian rhythms were observed in virtually all DOC patients. Most obvious, the very cyclicity of sleep and wakefulness, i.e. the typical distribution of sleep over the day and night, was severely disturbed. Although there is some evidence that the disturbance may root in a damage or severe dysfunction of the brain stem [40], purely cortical dysfunctions can also yield similar results, particularly when they are broadly spread like the diffuse axonal injury or diffuse gray matter damage after brain hypoxia. Even MCS patients slept less in the night and more during daytime than control patients. Much stronger was this anomaly in UWS patients in which the distribution of sleep and wakefulness did not differ between day and night. Some of the UWS patients did not show night sleep at all but slept only during the day. This observation underlines the necessity of polysomnographic recordings for at least 24 hours, because the data of only night sleep can be strongly misleading. Given that a recording day might occasionally be atypical, data collection for a still longer period (48 or 72 hours) would be useful [41, 42], but not at the cost of the decrease of sample size [43].

The presence of numerous sleep episodes during the daytime may account for strong fluctuations observed in the diagnostic behavioral assessment of both MCS [6] and UWS [8]. It is expected that by determining the distribution of periods of decreased arousal level, the risk of diagnostic misjudgment can be reduced.

Although the correlation between behavioral sleep (eyes open-/-closed) and electrophysiological (polysomnographic) signs of sleep was highly significant, it strongly varied among the groups. The correlation was close to 1.0 in control patients, and significantly decreased with the severity of DOC, indicating the possibility of behavioral/electrophysiological sleep dissociation in DOC patients. This finding supports the claim that from the appearance of eyes-open/-closed periods in DOC patients, it cannot be directly concluded that the patient is awake or asleep [2].

Whereas all sleep stages were present in all control patients, some sleep stages were entirely lacking in DOC patients. Particularly, significantly fewer UWS patients showed any sign of REM sleep than MCS and control patients. This finding is in line with previous studies, observing greater REM sleep anomalies in UWS than in MCS patients [10, 18, 40, 44]. Other studies reported lower amounts of REM sleep in DOC compared with healthy participants [45]. It has been suggested that disturbances of REM sleep indicate brain stem damage and could thus improve the identification of lesions in neuronal tissues and inform more targeted treatment of individual patients [46]. It could even be speculated that there is a specific relation between REM sleep and consciousness: REM sleep deficits might be associated with a lack of dreams, with dreams in turn being peculiar sleep states of consciousness. Furthermore, REM sleep has been proposed to serve the function of preparing the brain for the following state of wakefulness (evidence for this point was reviewed in [47]). It is, however, much too early to speculate about possible causal relationships.

In the present study, DOC patients also showed marked abnormalities with regard to sleep spindles. A total of 15 patients did not show any sleep spindles in S2. Moreover, even the DOC patients who did have idetifiable spindles, displayed an overall smaller number of spindles and a lower spindle amplitude than control patients. This finding is in line with previous studies that reported sleep spindle deficits in DOC patients [20, 40] and greater abnormalities in UWS than in MCS in terms of sleep spindles [44]. Spindle density is generally reduced in severe impairments of cognition and consciousness such as dementia [48, 49] and schizophrenia [50, 51]. But to the best of our knowledge, there are no clinical conditions where sleep spindles are completely absent. This absence may indicate a loss of thalamic circuits integrity [52], which presumably plays a role in the development of DOC [53].

On the background of these multiple abnormalities, there are a number of sleep features for which we did not observe differences between DOC and control patients. The total amount of sleep in control patients was similar to that reported in completely healthy individuals. The amount of sleep in DOC patients was slightly decreased, but this decrease did not reach significance. The average amount of S2 appears to differ between patient groups (CC > MCS > UWS), but the intragroup variance in S2 was so large (see Table 2) that the intergroup effect did not become significant. The amount of SWS was virtually identical in all three groups and, moreover, identical to the amount of SWS in normal populations of the corresponding age [25]. In this case, the F-ratio of 0.1 and BF of 5.29 indicate that the lack of significance may be a real null effect and not just a product of insufficient power. If this is true, this finding may indicate that SWS is so important as to maintain its overall stability even under conditions of severe distortions of consciousness and notwithstanding strong fluctuations in the distribution between day and night.

Patients with brain hypoxia were found to stay longer in sleep stage S1 than patients with other etiologies, while S2, SWS, and REM were unrelated to the cause of the DOC. Such a specific effect on S1 was unexpected because scoring of S1 is particularly difficult even in healthy sleepers. A possible explanation might be, e.g., that a poorly formed, difficult to assess pattern of background wakefulness EEG was observed in 70% of hypoxic patients but only in 23% of other patients. Generally, it is easy to confuse S1 with wakefulness state, and that such mistakes may be expected to be more frequent in patients with worse wakefulness EEG than in patients with well-formed wakefulness EEG. However, we would suggest to wait with a definite interpretation of the result until it has been replicated.

While our findings are largely in line with previous data, there are some differences. In contrast with our results, a large study by Rossi Sebastiano et al. [12] found a significant difference between UWS and MCS in terms of the presence of SWS but not of REM sleep. In a smaller study, Bedini et al. [54] did not observe any differences between UWS and MCS neither in SWS nor in REM sleep. There are at least two important differences between our study and these previous studies. First, it is not quite clear how sleep stages were scored in these previous studies and whether the raters were blind with regard to the patients’ diagnosis. Perhaps even more important, Bedini et al. and Rossi Sebastiano et al. performed polysomnographic recordings for less than 24h. As we have seen that DOC patients, and particularly UWS patients, can show any sleep stage at any time of day/night, it is quite possible that some important sleep epochs were missed during those 5 to 8 hours when the recording was not performed.

A big difficulty in the interpretation of sleep abnormalities in DOC is that the life conditions of such patients differ drastically from those of healthy individuals with respect to many factors, each of which can potentially disturb sleep. DOC patients spend most of their time in bed; during the night they are regularly disturbed by the light and sounds that cannot be completely avoided in a hospital setting; they are periodically awaked and moved by the personnel to avoid decubitus. Finally, they do not experience the usual social pressure to stay awake during daytime, and the fact that they can sleep almost whenever they want naturally decreases their need in night sleep. These are only a few of numerous external factors of disturbed sleep besides the internal factors related to brain lesion.

Independent studies demonstrated considerable effects of such environmental factors on REM sleep in intensive care unit patients [55, 56], but similar studies in DOC patients are entirely lacking. The present study was the first to include a clinical control group that was exposed to the same external environmental factors as the examined DOC patients, but did not suffer from conditions of brain damage. Even though this group contained only ten patients, their sleep pattern was radically different from that of DOC patients. Therefore, we conclude that the external factors mentioned above only play a minor, if any, role in the development of massive sleep abnormalities characteristic for DOC.

The conclusions drawn from the present results are, of course, limited by the moderate size of the sample. Some negative results (e.g., the non-significant findings in the domains of total amount of sleep and the amount of stage 2) might be different in a double or triple sample. Similarly, our sample size did not permit us to perform subgroup analyses, e.g., a comparison of MCS+ versus MCS-. Large multi-center studies would be necessary to investigate such effects, although serious organizational issues remain.

Another limitation, already mentioned above, is that 24 h is the minimal, but not the optimal recording time. Therefore, an atypical day might eventually have been caught in some patients. Again, however, 48 or 72 h polysomnographic recordings would present a considerable economical and technical challenge.

We investigated 24h distribution of neurophysiological and behavioral correlates of sleep in DOC patients. However, sleep is a complex physiological phenomenon and a truly comprehensive analysis of patients’ circadian rhythms would have to include also peripheral measures such as body temperature, melatonin and cortisol level.

Like other neurophysiological approaches (e.g., based on EEG, ERP, fMRI, or PET), polysomnography in DOC is still at the stage of investigation and cannot be directly applied as a diagnostic test. The situation is complicated by the lack of error-free golden standard with which a neurophysiological method can be matched. Large prognostic studies should be conducted to evaluate usefulness of sleep patterns in prediction of the clinical outcome, rather than in differentiating the clinical diagnosis. Another potentially important application of sleep studies can be identification of the individual time window of optimal wakefulness: the optimal time when clinical and electrophysiological assessment should be carried out.

## Conclusions

Our study indicates that the distribution of sleep signs in DOC patients over 24 hours differs significantly from the normal sleep-wakefulness pattern. Specifically, first, DOC patients show severely disturbed circadian rhythms with equally probable sleep occurrence during the day and night in UWS patients; second, eyes-closing behavior is less strongly correlated with neurophysiological sleep patterns; i.e., DOC patients spend longer time with eyes closed while awake than other neurological patients; third, some DOC patients do not have sleep spindles and REM sleep. Notably, these abnormalities of sleep in DOC are due to their brain lesion and cannot be attributed to external factors such as severe immobility and hospital environment.

## Data Availability

The data will be made freely available upon acceptance at a journal.

## Declarations

## Acknowledgements

We thank the project coordinator of the Schoen Clinics for Neurological Rehabilitation Barbara Schäpers, the nurses who collaborated with the scientists, and the patients’ families.

## Consent for Publication

Not applicable.

## Availability of data and materials

The datasets used and/or analyzed during the current study are available from the corresponding author on request.

## Author’ Contributions

BK, IM, YGP, FM, and SD contributed to the conception and design of the study. IM collected the data. YGP, IM and CB scored the sleep data, supervised by SD. YGP analyzed the data and prepared the figures. IM contributed to the data analysis. IM and YGP drafted the manuscript. BK, FM, CB, and SD brought major revisions in significant proportions of the manuscript.

All authors read and approved the final manuscript.

## Ethics approval and consent to participate

The study was conducted in accordance with the ethical standards of the Declaration of Helsinki and was approved by the Ethics Committee of the Faculty of Medicine, University of Tübingen. The patients and their legal representatives were informed about the study content before study enrollment and gave their written consent.

